# HLA Class I and Plasma Viral Load of HIV-1 in Sexually Transmitted and Reproductive Tract Infections among Heterosexual Serodiscordant couples in Nigeria

**DOI:** 10.1101/2021.08.23.21262283

**Authors:** NM Otuonye, Luo Ma, Chris Chinweokwu, MN Aniedobe, RN Okoye, VN Enya, FN Ogbonna, R Audu, M Uwandu, A Adedeji, J Ponmak, S Nduaga, SK Akindele, GO Liboro, EO Odewale, AA Adesesan, AZ Musa, O Ezechi, MM Ojetunde, NN Odunukwe

## Abstract

**Background:** This study investigated HLA Class I in Long Term Non-progressors (LTNPs) and plasma viral load in Sexually Transmitted and Reproductive Tract Infections (STIs/RTIs) associated with Heterosexual HIV-1 transmission among serodiscordant couples in Nigeria.

**Methods:** A total of 271 serodiscordant and concordant couples (HIV positive and negative) were enrolled, blood samples were collected from the subjects by venipuncture. HLA class I (with specific primers), plasma viral load, CD4+ analysis was done. Endocervical/urethral swabs and early morning urine samples were collected by standard microbiological methods. These were screened by microscopy, culture, antibiogram, and biochemical tests with a view to identify aetiologic agents of co-infections with HIV.

**Results:** The Participants age ranged from ≥ 21- < 50years. The index whose plasma viral loads were 10,001-100,000 copies/ml had STIs/RTIs 32(60.9% p=0.059). *Staphylococcus aureus and Escherichia coli* (22.1%) were isolated from the index (HIV positive subject) while 14.5% of Staphylococcus aureus and 27.2% of *E coli* were isolated from their partners (HIV negative subject). *Staphylococcus aureus* and *E coli* are normal flora but because the patients are Immunocompromised as a result of positivity to HIV, *Staphylococcus aureus* and *E. coli* in this context becomes opportunistic thereby, causing genital tract infections. *Staphylococcus* from the index showed more sensitivity to Amoxicillin/clavulanate (95.4%/90.4%) compared to the partners (55.1%/73.5%) and more resistant to Ceftazidime (81.4%) compared to the partners (68.9%). LTNPs were 28(8.51%) among the index. HLA-B alleles: B*5701 (9.2%), B*5703 (4.6%) and B*5801(12.5%) were identified for viral control at late stage of HIV infection while A*1 (4.6%), and C*0701 (29.1%) were protective alleles observed. HLA-B*0702 (33.3%), B*4201/A*2301(4.6%) respectively were susceptible alleles associated with seroconversion among LTNPs.

**Conclusion:** The microorganisms isolated from the index were associated with high viral loads and are independent makers to HIV-1 transmission among serodiscordant couples. Individuals associated with HLA class I alleles identified among LTNPs were those significantly associated with resistance and susceptible to HIV-1 infections.

## INTRODUCTION

About 36.7 million people globally are currently living with HIV infection^1, 2^. Nigeria has the 2nd largest HIV epidemic in the world with prevalence of 3.1% and has the highest new infection rates in sub Saharan Africa^3^. HIV is a public health problem; over 65% of the world’s HIV infection is found in sub Saharan Africa with heterosexual exposure as the primary mode of HIV transmission^4^. Approximately 80 percent of HIV infections in Nigeria are as a result of heterosexual sex^4^. However, many couples do not know their HIV status and so, this has given room to most heterosexual HIV-1 transmission among serodiscordant couples^3, 4^. Again, an estimated 70% HIV -1 transmission occur between married partners, making cohabiting African couples the largest HIV risk group^5, 6^. Factors contributing to this includes: Lack of knowledge about sexual health, non to inconsistent condom use, untreated ulcerative and non-ulcerative sexually transmitted diseases, reproductive tract infections, non-disclosure of partner HIV status, multiple sexual partners and vaginal washing^3, 7, 8^. However, gender inequality among women has been identified as a strong factor to HIV epidemic in African women^8^.

Vaginal infections which are caused by bacterial vaginosis (BV), bacterial pathogens (BP) and yeast infections (YI) are common among HIV-infected men and women especially during the reproductive age. This may be a marker for increased transmissibility to sexual partners, infants at delivery, significant morbidity, underscoring their importance from a public health perspective. Severe co- infections have been reported in serodiscordant couples. In a recent study, 564 patients presented to the Nigerian Institute of Medical Research (NIMR) HIV clinic with symptoms of lower waist pain, painful urination, vaginal/ urethral itching, discharge, certain foul-smelling fluid from the vagina and/or a balloon-like feeling from the vagina. Results presented showed fifty-five bacterial isolates co-infected with Candida species, while 5 *Trichomonas vaginalis* co-infected with candida species, 23 bacterial vaginosis (BV) co-infected with other bacterial pathogens^8^. About 10 patients had triple infection of BV, yeast and bacterial pathogens. Co-existence of reproductive tract infections (RTIs) was associated with increased HIV transmission through heterosexual contact (p<0.002)^8^. Though, traditionally, reproductive tract infections have been viewed as a lesser public health importance. In another study on lower genital tract infections in HIV-seropositive women in India, the laboratory findings showed high prevalence of BV (30%), mixed infection (30%), and candidiasis (10%) among HIV-seropositive women *(P* < 0.001)^9^.

Another study in India identified sexually transmitted infections in 57% of HIV positive women compared to 34% of HIV negative women (p=0.0037). Vaginal candidiasis was the most common infection followed by *Trichomonas vaginalis.* Human papilloma virus infection was seen in nine HIV positive women and none was seen in HIV negative women^10^. Symptomatic vaginal yeast infections are increased in HIV-infected women with low CD4 cell count if they are not on antifungal prophylaxis. Significantly, vaginal infections may cause significant morbidity, especially among HIV-infected women, and may contribute to increased risk of HIV transmission among sexual partners. Serodiscordant couples involved in oral sexual practices can be of high risk in HIV-1 transmission if there are open sores in the mouth. The sexual practices with the highest risks are those that cause mucosal trauma, during intercourse while anal- receptive intercourse poses the highest risk. Mucous membrane inflammation facilitates HIV transmission. Sexually Transmitted Diseases, such as gonorrhea, chlamydial infection, trichomoniasis, cause ulceration. Chancroid, herpes, and syphilis, increase HIV risk acquisition in several folds. In heterosexuals, the estimated risk per coital act is about 1/1000^11^. However, the risk is increased in early and advanced stages of HIV infection when HIV concentrations in plasma and genital fluids are higher, in younger people with ulcerative genital diseases^12^. The risk of HIV acquisition in a single sexual exposure is estimated to be very low^13^. However, the key factor in determining the viral load burden is the exposing dose of the virus from an Index to the partner^14^. A study conducted in Uganda on heterosexual HIV serodiscordant couples showed a direct relationship between the risk of HIV acquisition and donor plasma viral load (pVL)^15^. No HIV transmissions were observed when the pVL was below a critical threshold of 1500 copies HIV RNA/ml ^15^.

Human Leucocyte Antigen (HLA) play a crucial role in HIV diseases progression or seroconversion^16, 17^. However, the role HLA play in HIV-1 transmission in serodiscordant couples in Nigeria is unknown. HLA is the human version of the Major Histocompatibility Complex in man^18^. HLAs are complex, genetically inherited proteins present on the surface of human cells, located at the short arm of chromosome 6 (6p21) and spans approximately 4.0 kilobases of DNA. It is known to be the most polymorphic genetic system in humans^16^. HLA is made up of class I (HLA-A, -B and -C) and class II (DP, DQ, DR, DM, DP and DP) ^17^. Several studies particularly on Western populations have demonstrated their implications in many autoimmune and infectious diseases including HIV^17^. The biological role of the HLA class I and II molecules are to present processed peptide antigens (HIV, bacteria and other viruses) to CD8+ cytotoxic T lymphocytes and CD4+ respectively for immune recognition and stimulation with the aim of destroying the pathogen from which the antigenic peptide came from^18^. Studies have shown that, the genetic make-up of a person’s HLA affects the rate of HIV disease progression^16^.

Long Term Non-progressors are a minor percentage of individuals (5–10%) who remains asymptomatic of HIV infection^19^. These subset of HIV-1 infected individuals can control HIV replication which is linked to the expression of certain HLA class I alleles particularly B*5701, B*5703, B*27 and B*5801^20^. LTNPs’ immunologic basis is also linked to cytotoxic lymphocyte functions (CD8+). Their CD4^+^ T-cell counts is >500 cells mm^−3^ for >10 or more years with viral loads under 1,000 copies RNA/ml blood^21^. Many of these patients have been HIV positive for 30 years without ARV medications^21^. Some authors suggested that some LTNPs are infected with a weakened or inactive form of HIV while some believe that many LTNP patients carry a fully virulent form of the virus^19^. Again, some HLA alleles have been documented by another study to determine the rate of HIV-1 seroconversion and disease progression in 1000 Pumwani sex workers in Kenya^22^. From their analysis, they reported that women with HLA- A*01, C*0602 and C*0701 are less likely to seroconvert and conversely women who have A*2301 (*P* = 0.004), B*070201 (*P* = 0.003) and B*4201 (*P* = 0.025) are independently associated with rapid seroconversion. Therefore, it becomes necessary to know if these LTNPs also harbour some of these HLA alleles previously documented to protect, restrict or lead to HIV-1 disease progression.

In this study, we therefore;

1. Investigated the role of plasma viral load (HIV-1 RNA) in association with sexually transmitted infections and reproductive tract infections (STIs/RTIs) and behavioural co- factors in HIV-1 transmission among serodiscordant couples.
2. Identified the antibiotic susceptibility and resistance patterns of the STIs/RTIs isolates predominant in serodiscordant couples.
3. Documented the profile of HLA class I alleles in Long Term Non-Progressors in association of plasma viral load in transmitting HIV -1 from index to their partners
4. Matched specific HLA alleles in Long Term Non- Progressors associated with protective, resistance and susceptible to seroconversion to HIV diseases with other alleles identified in previous studies.

## Subjects, Materials and Methods

### Recruitment of study population

The study population comprised of: Serodiscordant couples (Index and Partner). The positive partner may or may not be on antiretroviral drugs (ART). Concordant couples were also recruited: in this case, the couple can be concordant HIV positive and or concordant HIV negative. These couples were registered with the Nigerian Institute of Medical Research (NIMR) HIV clinic Yaba, Lagos and Nnamdi Azikiwe University Teaching Hospital Nnewi Anambra State (NAUTH) HIV clinics. Other clients were recruited from NIMR HCT Unit (HIV Counseling and Testing Unit). HIV negative couples were recruited among NIMR Workers (NIMRWC) and Divine Grace Church Couples (DGCC) for control measures. Only couples who gave informed consent and were recruited.

### Study design

Case–control study of the people living with HIV/AIDS and their HIV negative partners (serodiscordant couples), concordant HIV positive couples and concordant HIV negative couples (control) with previous or current history of cervicitis, pelvic inflammatory disease (PID), painful urination, itching and foul-smelling vagina, were enrolled for the study.

### Focus group discussion and Consent documentation

Participants were counseled and told to inform their partners about the research in other to facilitate their consent. The partners of all index were screened for HIV. Those who subsequently turned out to be HIV positive were referred to NIMR and NAUTH HIV Counseling and Testing unit for confirmation of their sero-positive status, and finally referred to the HIV clinic for management. Couples recruited from amongst NIMR workers (NIMRWC) and Divine Grace Church Couples (DGCC) were all screened for HIV to confirm their sero-negative status. Informed consent was obtained from all the participants. This was obtained during focus group discussions conducted in an appropriate meeting room.

### Study Sites

Samples were collected and processed in two government-owned health institutions namely: Nigerian Institute of Medical Research Yaba, Lagos State and Nnamdi Azikiwe University Teaching Hospital Nnewi, Anambra State. HLA studies were carried out at National Microbiology Laboratory Winnipeg, Manitoba Canada.

#### Sample size Calculation

This N=Z^2^ Q (1-P)/ D^2^ was used with HIV prevalence of 3.4% a sample size of 50 was gotten as follows: N= 0.034 X3.8416 X0.966/ 0.0025=50.

Following the HIV prevalence rate used (3.4%), the sample size to be used for the study is 50 but considering how little the sample size was, I had to make it a case control study so that a larger population can be recruited in the study. The initial calculation in the first paper sent was wrong and that was why it was looking like an understudy.

### Inclusion and Exclusion Criteria

The participants included in this study were: (1) Couples that have discordant HIV test result. (2) Couples that have concordant HIV results (3) Couples who have co-habited for at least 3 months and or may have more than one partner. (4) The women are aged between 20-60 and the men 20-65 years. (5) The HIV positive partners can be or not be on ARV. (6) Willingness to participate and given informed consent.

Also excluded were (1) Those over/or under the required age group (2). Those, who have not co- habited for three months (3) Those who are not willing to participate and give informed consent.

### Ethical Approval

Approval was obtained from NIMR IRB (IRB/12/176) and NAUTH Ethical Committee (CS/66/7/79).

### Administration of Structured Questionnaire using In-depth Interview (IDI) Procedures and face to face interviews

Structured questionnaires were administered to a total of 224 consenting sero-discordant couples, 26 concordant HIV positive and 21 concordant HIV negative couples. Information on socio- demographic characteristics, knowledge of partners’ HIV status, knowledge of STIs, laboratory tests and treatment status, sexual behaviours and practices, were required for the completion of the questionnaire. This procedure was done during couples counseling and testing

### Couples Voluntary Counseling and Testing (CVCT) for HIV Prevention

In this study 99.8% of the couples accepted to be counseled together. The 0.2% that were counseled separately but later brought together was because their wives suspected their husbands of infidelity and gave room for their husbands to say the truth. However, counseling both partners in a serodiscordant relationship at the same time (couples-based counseling) has been proved to be more effective in reducing risk behaviours compared to counseling the partners individually^25^. In this study, couples-based counseling created a supportive space where partners came to an agreement on how to reduce their risk of HIV transmission and develop ways to support each other in using HIV prevention strategies continuously and correctly. Couples were supported to discuss potentially sensitive issues relevant to HIV prevention, such as sexual intimacy, interpersonal dynamics and whether there are sexual partners outside of the relationship. Through counseling, heterosexual serodiscordant couples explored if the plans to have children may influence the acceptance of HIV prevention strategies.

### Laboratory investigations Sample collection methods

A total of 472 blood samples, 123 endocervical swabs, 11 urethral swabs, 109 urine samples were collected from 236 couples from Nigerian Institute of Medical Research. Seventy (70) blood samples, 28 urine samples and 3 endocervical swabs were also collected from 35 couples from Nnamdi Azikiwe University Teaching Hospital Nnewi Anambra State. The participants that were swabbed and brought early morning urine samples presented with: vaginal discharge, pelvic inflammatory diseases (PID) itching, painful urination and foul smell from the vagina. The swabs and the urine samples were cultured using standard microbiological methods.

### Isolation and Identification of microorganisms associated with Sexually Transmitted and Reproductive Tract Infections

#### Culture

Endocervical/urethral swabs were cultured on chocolate and blood agar under (CO_2_), and Sabouraud Dextrose agar plates and placed at 37 ^0^C for 24 hours. Additionally, urine samples were cultured on MacConkey, CLED and blood agar plates, placed at 37^0^C for 24 hours, using standard microbiological techniques. Characterization and identification were done on the microbial isolates using standard microbiological techniques such as cultural, morphological and biochemical characteristics accordingly ^26^.

### Wet Preparation

This was done on endocervical swabs and urine samples to determine the presence of clue and yeast cells and *T. vaginalis* using standard microbiological methods ^25^.

#### *Gardnerella vaginalis* detection (Bacterial Vaginosis {BV})

BV diagnosis was done using the presence of the three Amstel criteria which include vaginal pH, greater than 4.5, positive whiff test, milky discharge, and the presence of clue cells on microscopic examination of vaginal fluid^27^.

### Antibiotic Susceptibility Testing

The disk-diffusion agar method tests the effectiveness of antibiotics on a specific microorganism. The inoculum for each bacterial isolate was prepared directly from corresponding overnight culture after 24 hours incubation at 37°C were prepared directly from overnight cultures adjusted to 0.5 McFarland standard. Whatman Antibiotic Assay Single Discs (9mm) already impregnated with a known volume of appropriate concentration of antimicrobial, representing Gram positive and negative antibiotics compromising of: Ofloxacin (5µg), Gentamycin (10µg), Amoxicillin/Clavulanate (10µg), Amoxicillin (10µg), Ciprofloxacin (5µg), Nitrofurantoin (100µg) , Erythromycin (10µg), Cefotaxime (30µg), were placed aseptically on the inoculated plates. The plates were incubated for 24 hours at 37^0^C. Clear Zones of Inhibition was measured in millimetres using a ruler on the underside of the plate. Zone measurements (+/- 2 mm) with 96% of the tests was accepted as the right zone of inhibition according to the Kirby-Bauer procedure^28^. Isolates showing resistance to three or more categories of antibiotics were considered as multidrug resistance bacteria.

### HIV-1 RNA Assay (plasma viral load)

Determination of the Amplicor HIV-1 MONITOR Test version 1.5 is an in vitro nucleic acid amplification test for the quantitation of RNA copies/ml of plasma of HIV-1 RNA in human plasma. Five stages were involved: Pre-PCR reagent, Pre–PCR standard specimen preparations, amplification, detection, calculation and interpretation of results. The HIV-1 RNA copies/ml of plasma were calculated as copies/PCR x 40 = HIV-1 RNA copies/mL ^29^.

### CD4 Count

Twenty (20µl) microlitre of whole blood, 20µl of CD4 mAb PE was added to Partec test tubes and incubated for 15 mins in the dark. Subsequently, 800 µl of no lyse buffer was added and vortexed gently. Total counting of CD4+ was done by Partec devise Cyflow which is a desktop flow cytometer. Result was displayed on the computer screen and calculated in cells/mm^3^. Normal range in HIV negative individual is 400-1600cells/mm3 as described^30^.

### White cell Harvesting and DNA Extraction

White cell pellet was harvested from buffy coat on peripheral blood cells with the use of Ace- shocking solution. The white cells pellet was used for Qiagen DNA extraction. DNA was extracted from white cell pellet with genomic DNAQIAGEN kit as described^31^. The DNA filtrate was placed in the cryovial and stored frozen at -800C or -200C for further analysis.

### Polymerase Chain Reaction (PCR) for DNA Amplification

PCR reactions were performed on 96-well plates. Each reaction was composed of 20µl of ddH_2_O, 5µl of gDNA, and 25µl of master mix. The master mix contained 22.75µl of 2X mix

(120mM Tris-HCl, 3mM MgCl_2_, 30mM (NH_4_)_2_SO_4_, 200µM dNTPs, 0.2% gelatin, ddH_2_O), 55 pmol of both the forward and reverse primer and 13.75 µl of Taq polymerase. For HLA-C typing, primers were set to detect polymorphisms in exons 2 and 3 of the class 1 genes. The structural sequence of the primers used for HLA-A, -B and -C were as follows: APCRF represents HLA-A Forward Primers and APCRF: HLA-A Forward Primers. BPCRR represents HLA-B Reverse Primers and BPCRF: HLA-B Forward Primers. While CPCRR represents HLA-C Reverse Primers and CPCRF: HLA-C Forward Primers as shown below ^32^.

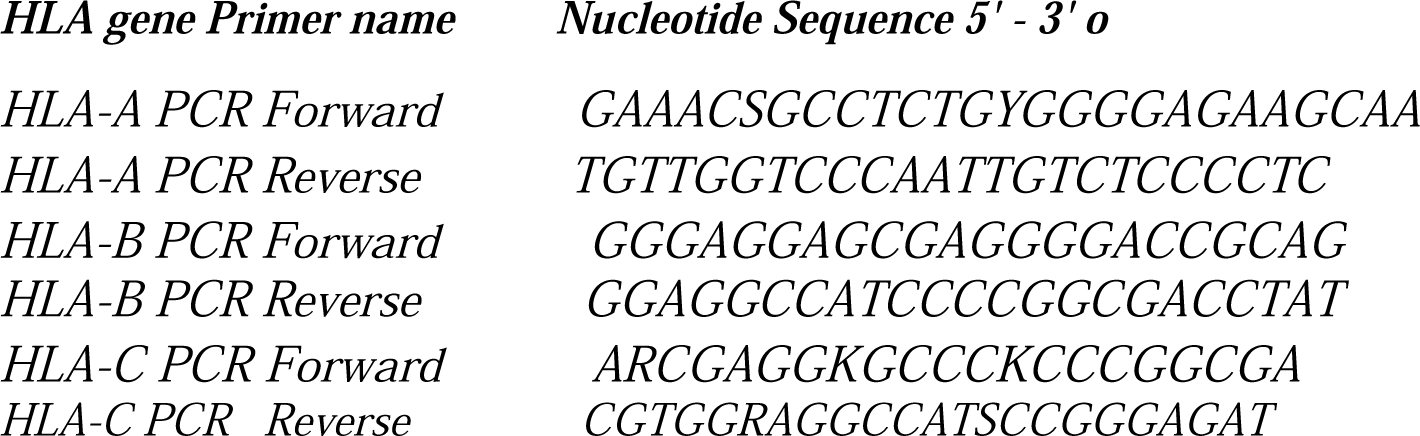

### Agarose gel Electrophoresis

This was used to confirm that PCR was successful in amplifying the desired HLA gene. Each sample as was run in a 1% (w/v) agarose gel in 1X TBE buffer and stained with 0.003% 10mg/ml Ethidium Bromide. A 1kb ladder was added to the first well of every row. Once the DNA had migrated to a sufficient distance down the gel, they were removed from the tank and illuminated using a BioRad™ UV Transilluminator 2000. The results were viewed with the BioRad™ Quantity One® software accordingly ^33^

### Purification of PCR Product

Following sequencing PCR, the PCR product was precipitated using 21µl of a mixture containing 5ml of 95% ethanol and 250µl of sodium acetate. To remove any buffers, salts and reagents. Finally, the contents of each well were transferred to MicroAMP™ plates and put in the Applied Biosystems™ 3130xl Genetic Analyzer for electrophoresis according to Agencourt® AMPure Protocol^34^.

### Sequencing HLA PCR

This protocol was done after the PCR products have been purified. Sequencing HLA is usually focused on the most polymorphic exons 2 and 3 which encode the antigen recognition site as described by Sanger ^35^. To genotype HLA class I alleles, exons 2 and 3 were sequenced^36^. Each sequencing PCR reaction contained 4µl of purified PCR product, 1.5µl primer at 3.2µM and 2µl of Applied Biosystems™. BigDye® Terminator V1.1 or V 3.1. Big Dye V 1.1 was used for HLA-B and -C and Big Dye V 3.1 for HLA-A. The Forward and Reverse Primers used for each HLA gene are shown on the table below:

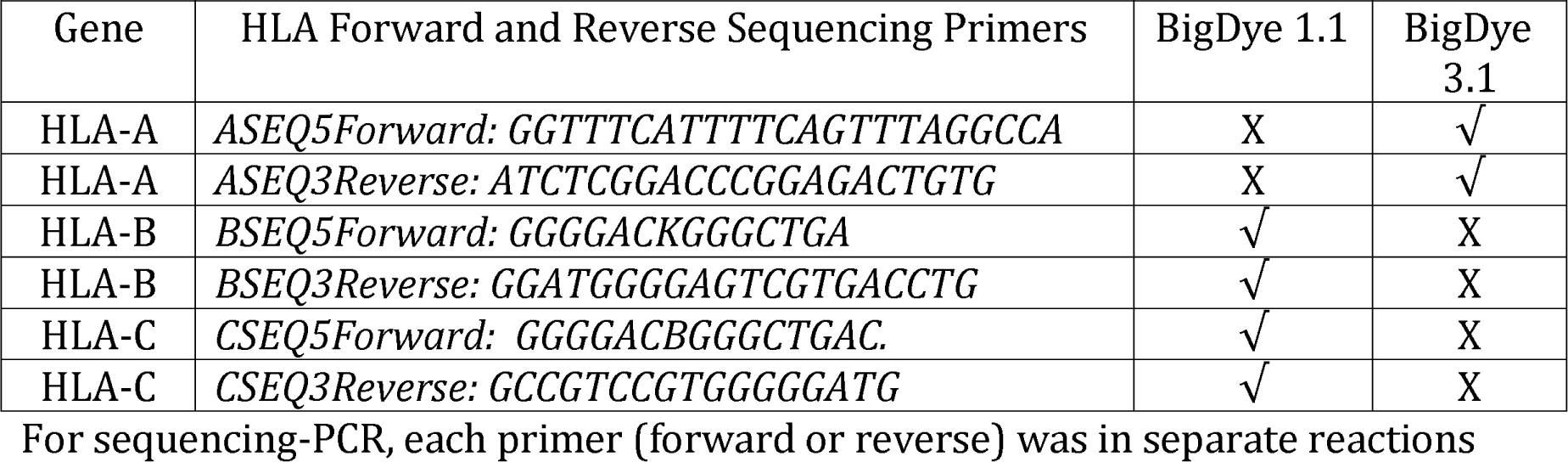

### Ethanol Precipitation

Following sequencing PCR, the PCR product was precipitated using 21µl of a mixture containing 5ml of 95% ethanol and 250µl of sodium acetate. This process removed any buffers, salts and reagents in the PCR product. Finally, the contents of each well were transferred to MicroAMP™ plates and put in the Applied Biosystems™ 3130xl Genetic Analyzer for electrophoresis.

### Biosystems™ 3130xl Genetic Electropherogram

Each respondent’s sequenced PCR products were loaded in the Biosystems for electrophoresis and data analysis. Data analysis software processed the raw data in the *ab1 file, using algorithms which applies the Multicomponent analysis that separates the four different fluorescent dye signals into distinct spectral components by mathematically filtering fluorescence signal from dyes with emission spectra overlap Basecalling. The selected Basecaller processes the fluorescent signals then assigns a base to each peak (A, C, G, T, or N). Analyzed sample data was displayed as an electropherogram, a sequence of peaks in four colors.

Each color represents the base called for the peak^37^

### CondonExpress TM

CodonExpress is a genotyping software based on a Taxonomy-based Sequence Analysis (TBSA) to resolve HLA Heterozygous and homozygous combinations. CodonExpress was used to type the Sequence-Based HLA class 1 genes according to CodonExpress TM, as described by the University of Manitoba, Canada instruction manual^38^.

### Sequencher V5 for Data Integrity

Data Integrity was done by using DNA Sequencher V5 Software. This was done to ensure that all data generated in this study are correct and to rule out all unresolved HLA typing ambiguities as previously reported^39^.

### Statistical Analysis

Data generated was entered into IBM SPSS statistics version 20. Cross-sectional analysis using Chi Square was used to identify association between genetic and non-genetic factors. Confidence interval was set at 95% with level of significance set at *p* value less than 0.05 (*P* < 0.05)^40^.

## Results

### The sociodemographic characteristics

The sociodemographic characteristics of the 271 (542) study participants by site of recruitment are shown on table 1. Their ages ranged from 20 to 60 years with median age of 38 years. Sixteen couples 16 (3.0%) out of 271 couples were engaged to be married but have been co- habiting and are serodiscordant. The ethnicity showed that 54.9% were Igbos, 20.9% Yoruba, 9.3% Hausa and 14.9% were of other ethnic extractions. HIV status showed 224 (82.7%) serodiscordant, 26(9.6%) concordant HIV positive and 21(7.7%) concordant HIV negative couples as shown on table 1.

**Table 1:**
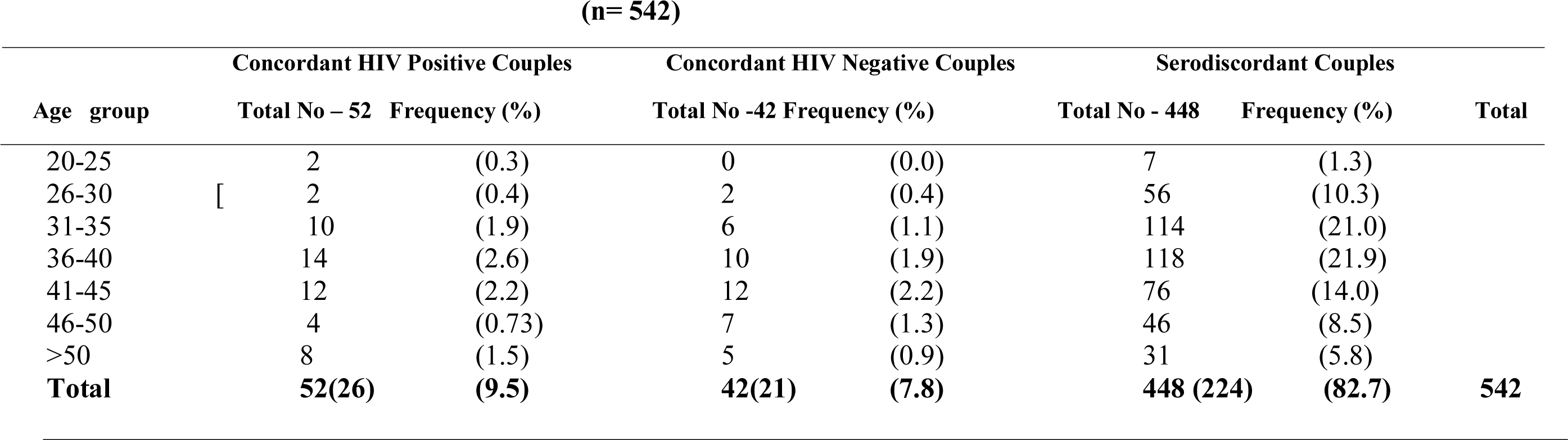
Sociodemographic Characteristics of Study Participant

**(n= 542)**
| Age group | Concordant HIV Positive Couples |  | Concordant HIV Negative Couples |  | Serodiscordant Couples |  | Total |
| --- | --- | --- | --- | --- | --- | --- | --- |
|  | Total No – 52 | Frequency (%) | Total No -42 | Frequency (%) | Total No - 448 | Frequency (%) |  |
| 20-25 | 2 | (0.3) | 0 | (0.0) | 7 | (1.3) |  |
| 26-30 | 2 | (0.4) | 2 | (0.4) | 56 | (10.3) |  |
| 31-35 | 10 | (1.9) | 6 | (1.1) | 114 | (21.0) |  |
| 36-40 | 14 | (2.6) | 10 | (1.9) | 118 | (21.9) |  |
| 41-45 | 12 | (2.2) | 12 | (2.2) | 76 | (14.0) |  |
| 46-50 | 4 | (0.73) | 7 | (1.3) | 46 | (8.5) |  |
| >50 | 8 | (1.5) | 5 | (0.9) | 31 | (5.8) |  |
| **Total** | **52(26)** | **(9.5)** | **42(21)** | **(7.8)** | **448 (224)** | **(82.7)** | **542** |

### The impact in HIV-1 RNA burden (plasma viral load (pVL) and other Biological and behavioural Risk factors

The impact in HIV-1 RNA burden (plasma viral load) on other risk factors on heterosexual HIV- 1 transmission is shown on table 2. The heterosexual index (*P* = 0.057) whose pVL was 10,001 to 100,000 had higher number of sexually transmitted and reproductive tract infections and showed statistical significance to sexually transmitted infections (STIs) and Reproductive Tract Infection (RTIs) (*P* = 0.059).

**Table 2:**
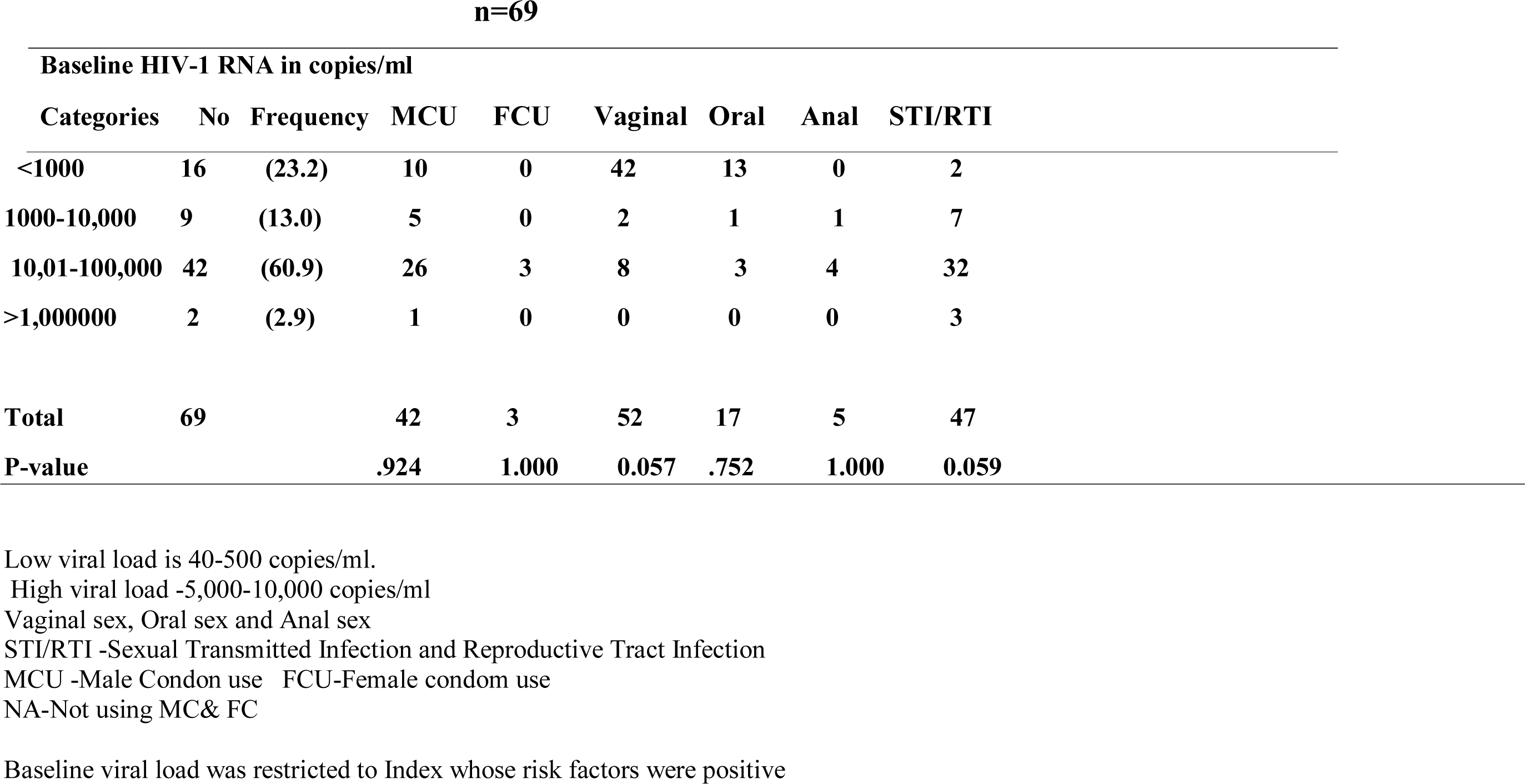
Impact of Baseline HIV-1 RNA (Viral load) and Other Risk Factors in HIV-1 Transmission in the Index

**n=69**
| **Baseline HIV-1 RNA in copies/ml** |  |  |  |  |  |  |  |  |
| --- | --- | --- | --- | --- | --- | --- | --- | --- |
| **Categories** | **No** | **Frequency** | **MCU** | **FCU** | **Vaginal** | **Oral** | **Anal** | **STI/RTI** |
| **<1000** | **16** | **(23.2)** | **10** | **0** | **42** | **13** | **0** | **2** |
| **1000-10,000** | **9** | **(13.0)** | **5** | **0** | **2** | **1** | **1** | **7** |
| **10,01-100,000** | **42** | **(60.9)** | **26** | **3** | **8** | **3** | **4** | **32** |
| **>1,000000** | **2** | **(2.9)** | **1** | **0** | **0** | **0** | **0** | **3** |
| **Total** | **69** |  | **42** | **3** | **52** | **17** | **5** | **47** |
| **P-value** |  |  | **.924** | **1.000** | **0.057** | **.752** | **1.000** | **0.059** |

Low viral load is 40-500 copies/ml.
High viral load -5,000-10,000 copies/ml
Vaginal sex, Oral sex and Anal sex
STI/RTI -Sexual Transmitted Infection and Reproductive Tract Infection
MCU -Male Condon use FCU-Female condom use
NA-Not using MC& FC
Baseline viral load was restricted to Index whose risk factors were positive

### Microorganisms isolated from people living with HIV (Index) and HIV negative (partners)

Most organisms isolated occurred as mixed infections in the population of HIV negative individuals which was shown on table 3a. A total number of 55 microorganisms were isolated and out of which, *Candida* spp. 17(30.9%) was the most prevalent followed by *Escherichia coli* 15(27.2%) and *Proteus mirabilis.* 2 (3.6%) being the least. *Escherichia coli* (11) and *Candida albicans* (11) mostly co-infected followed by *Staphylococcus aureus* (6) and the least is *Streptococcus* spp. (1). *Escherichia. coli* and *Candida* spp. co-infected with each other 5 and 7 times respectively. This is shown on table 3a.

**Table 3:**
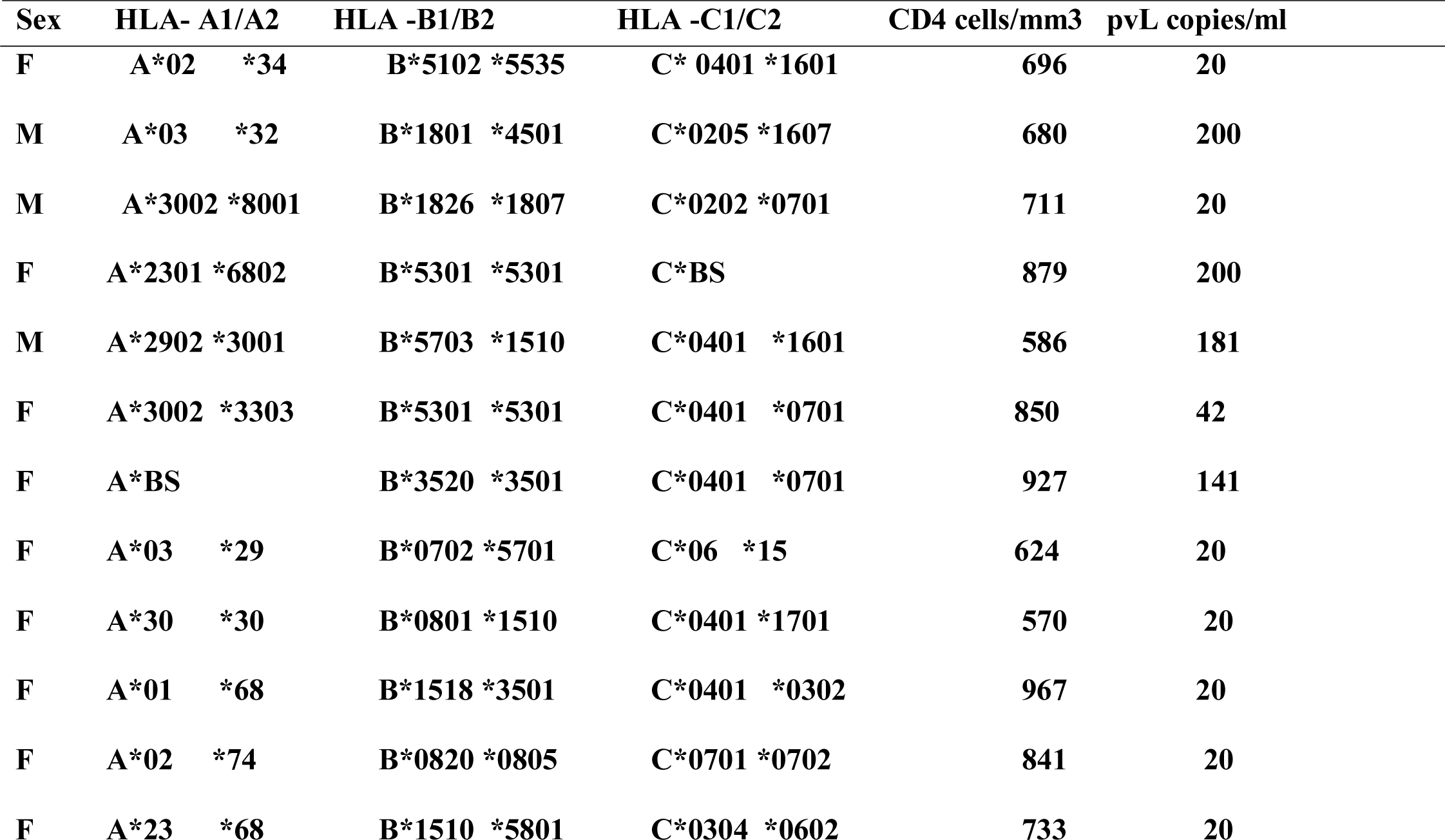
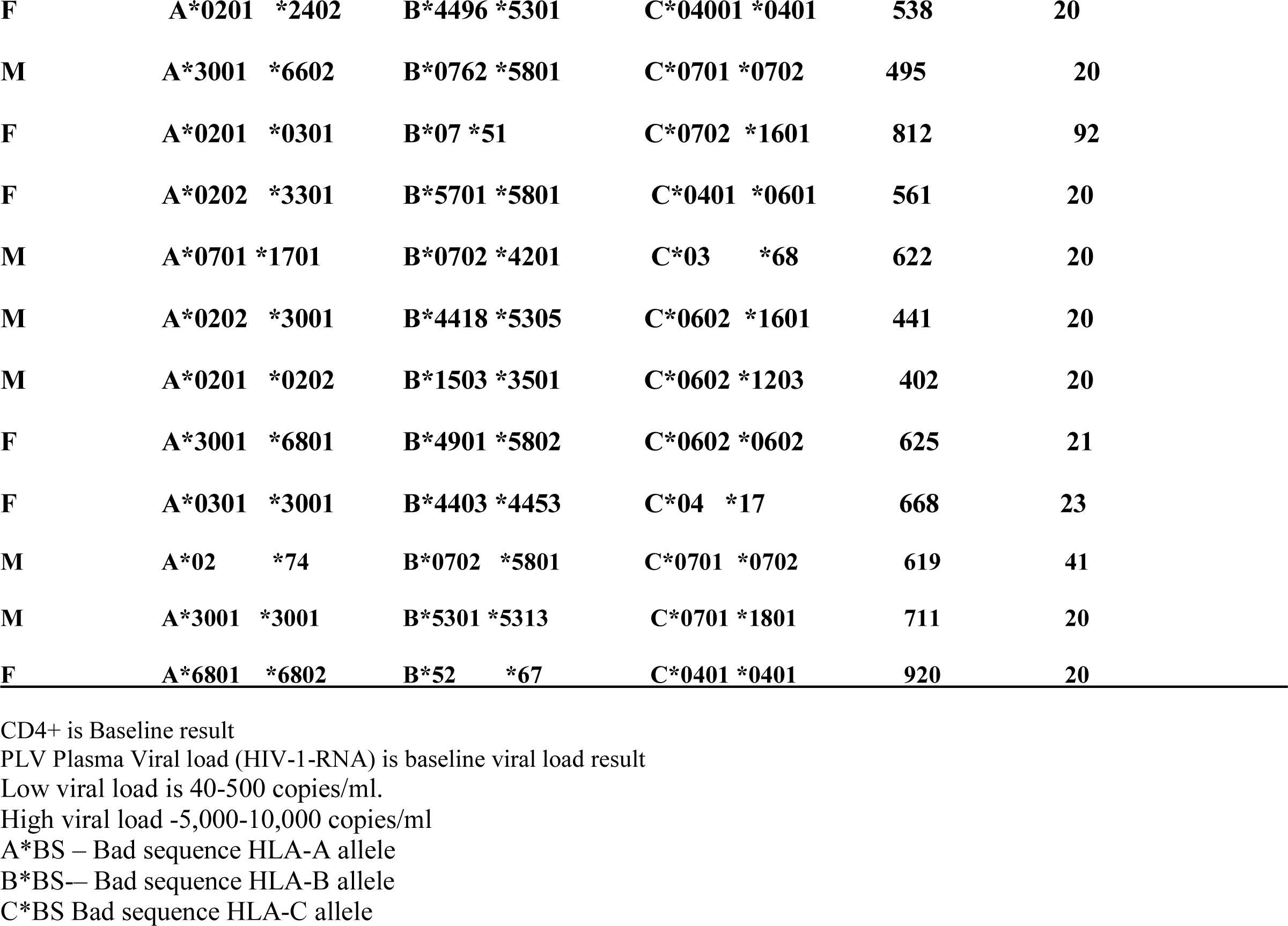
Profile of HLA-Class I Alleles CD4+ CD8+ and pVL of Long-term of Non -Progressor’s (LTNP) among HIV Positive Individuals

| **Sex** | **HLA- A1/A2** | **HLA -B1/B2** | **HLA -C1/C2** | **CD4 cells/mm3** | **pVL copies/ml** |
| --- | --- | --- | --- | --- | --- |
| **F** | **A\*02 \*34** | **B\*5102 \*5535** | **C\* 0401 \*1601** | **696** | **20** |
| **M** | **A\*03 \*32** | **B\*1801 \*4501** | **C\*0205 \*1607** | **680** | **200** |
| **M** | **A\*3002 \*8001** | **B\*1826 \*1807** | **C\*0202 \*0701** | **711** | **20** |
| **F** | **A\*2301 \*6802** | **B\*5301 \*5301** | **C\*BS** | **879** | **200** |
| **M** | **A\*2902 \*3001** | **B\*5703 \*1510** | **C\*0401 \*1601** | **586** | **181** |
| **F** | **A\*3002 \*3303** | **B\*5301 \*5301** | **C\*0401 \*0701** | **850** | **42** |
| **F** | **A\*BS** | **B\*3520 \*3501** | **C\*0401 \*0701** | **927** | **141** |
| **F** | **A\*03 \*29** | **B\*0702 \*5701** | **C\*06 \*15** | **624** | **20** |
| **F** | **A\*30 \*30** | **B\*0801 \*1510** | **C\*0401 \*1701** | **570** | **20** |
| **F** | **A\*01 \*68** | **B\*1518 \*3501** | **C\*0401 \*0302** | **967** | **20** |
| **F** | **A\*02 \*74** | **B\*0820 \*0805** | **C\*0701 \*0702** | **841** | **20** |
| **F** | **A\*23 \*68** | **B\*1510 \*5801** | **C\*0304 \*0602** | **733** | **20** |
| Page 2 of table 3 |  |  |  |  |  |
| **F** | **A\*0201 \*2402** | **B\*4496 \*5301** | **C\*04001 \*0401** | **538** | **20** |
| **M** | **A\*3001 \*6602** | **B\*0762 \*5801** | **C\*0701 \*0702** | **495** | **20** |
| **F** | **A\*0201 \*0301** | **B\*07 \*51** | **C\*0702 \*1601** | **812** | **92** |
| **F** | **A\*0202 \*3301** | **B\*5701 \*5801** | **C\*0401 \*0601** | **561** | **20** |
| **M** | **A\*0701 \*1701** | **B\*0702 \*4201** | **C\*03 \*68** | **622** | **20** |
| **M** | **A\*0202 \*3001** | **B\*4418 \*5305** | **C\*0602 \*1601** | **441** | **20** |
| **M** | **A\*0201 \*0202** | **B\*1503 \*3501** | **C\*0602 \*1203** | **402** | **20** |
| **F** | **A\*3001 \*6801** | **B\*4901 \*5802** | **C\*0602 \*0602** | **625** | **21** |
| **F** | **A\*0301 \*3001** | **B\*4403 \*4453** | **C\*04 \*17** | **668** | **23** |
| **M** | **A\*02 \*74** | **B\*0702 \*5801** | **C\*0701 \*0702** | **619** | **41** |
| **M** | **A\*3001 \*3001** | **B\*5301 \*5313** | **C\*0701 \*1801** | **711** | **20** |
| **F** | **A\*6801 \*6802** | **B\*52 \*67** | **C\*0401 \*0401** | **920** | **20** |

CD4+ is Baseline result
PLV Plasma Viral load (HIV-1-RNA) is baseline viral load result
Low viral load is 40-500 copies/ml.
High viral load -5,000-10,000 copies/ml
A\*BS – Bad sequence HLA-A allele
B\*BS-- Bad sequence HLA-B allele
C\*BS Bad sequence HLA-C allele

Table 3b highlighted the microorganisms isolated and the occurrence of mixed infections in HIV positive individuals. A total number of 208 microorganisms were isolated and *Candida albicans* 56 (26.9%) was the most prevalent followed by *Staphylococcus aureus and Escherichia. coli* 46 (22.1%) respectively while *Pseudomonas* aeruginosa had the least frequency 1(0.4%). *Trichomonas vaginalis*: (1.4%) were seen in HIV positive females. *Candida* spp. co-infected mostly with *Klebsiella* spp. (6) followed by *Staphylococcus aureus, Escherichia coli* and *Streptococcus* spp. (3) respectively. *Gardnerella vaginalis* co-infected with *Staphylococcus aureus* (4), followed by *Streptococcus* spp. and *Klebsiella* spp. (2) respectively.

### Antimicrobial Susceptibility Patterns of the Bacterial Isolates

On assessment of the antimicrobial sensitivity patterns of the bacterial isolates of reproductive tract infections in HIV positive and negative individuals, the following were observed: *Staphylococcus aureus* isolated from HIV positive individuals showed more sensitivity to Amoxicillin/clavulanate (95.4%) followed by gentamycin (90.4%) compared to HIV negative individuals (55.1%/34.4%). *Escherichia coli* isolated from HIV positive individuals showed more sensitivity to Nitrofurantoin followed by Gentamycin and Ofloxacin (95.4%/98.5%/98.0%) compared to HIV negative individuals (66.1%/73.5% /33.0%). *Klebsiella* spp. isolated from HIV positive individuals showed more sensitivity to Ofloxacin and Nitrofurantoin (87.7%) respectively compared to HIV negative individuals (27.7% /0.0%). This is shown on table 4a.

**Table 4a.** Aetiologic Agents Isolated and Occurrence of Mixed Infections in HIV Negative Individuals

| Aetiologic agents | Total No.<br>of Isolates (%) | CO -INFECTION WITH |  |  |  |  |  |  |  |  |
| --- | --- | --- | --- | --- | --- | --- | --- | --- | --- | --- |
|  |  | *S. aureus* | *Candida spp.* | *E. coli* | *B. vaginosis* | *T. vaginalis* | *Strept. spp.* | *Kleb. spp.* | *Proteus spp.* | *Pseudomonas spp.* |
| *S. aureus* | 8(14.5) | 2 | 1 | 2 | 0 | 0 | 0 | 0 | 0 | 0 |
| *Candida spp.* | 17(30.9) | 1 | 0 | 7 | 3 | 0 | 1 | 0 | 0 | 0 |
| *E. coli* | 15(27.2) | 1 | 5 | 0 | 1 | 0 | 0 | 0 | 0 | 0 |
| *G. vaginalis* | 3(5.5) | 1 | 1 | 2 | 0 | 0 | 0 | 0 | 0 | 0 |
| *T. vaginalis* | 0 (0.0) | 0 | 0 | 0 | 0 | 0 | 0 | 0 | 0 | 0 |
| *Streptococcus spp.* | 3(5.5) | 0 | 1 | 0 | 1 | 0 | 0 | 0 | 0 | 0 |
| *Klebsiella spp.* | 4(7.2) | 0 | 0 | 0 | 0 | 0 | 0 | 0 | 0 | 0 |
| *Proteus spp.* | 2(3.6) | 0 | 1 | 0 | 0 | 0 | 0 | 0 | 0 | 0 |
| *Pseudomonas spp.* | 3 (5.5) | 1 | 2 | 0 | 0 | 0 | 0 | 0 | 0 | 0 |
| **Total** | **55(100)** | **6** | **11** | **11** | **5** | **0** | **1** | **0** | **0** | **0** |

A total number of 55 aetiologic agents were isolated from HIV negative individuals.

On assessment of the antimicrobial resistant patterns of the bacterial isolates of reproductive tract infections in HIV positive and negative individuals, the following were observed: *Staphylococcus aureus* isolated from HIV positive individuals showed more resistance to Ceftazidime and Gentamycin (81.4%/ 72.3%) compared to HIV negative individuals (68.9%/6.8%). *Escherichia coli* isolated from in HIV positive individuals showed more resistance to Tetracycline and Ofloxacin (85.9%/76.9%) compared to HIV negative individuals (11.0%/33.0%). *Proteus* spp. isolated from HIV negative individuals showed more resistance to Gentamycin, Amoxil, Amoxicillin/clavulanate and Ceftazidime (83.3%) respectively compared HIV positive individuals (0.0%/48.0%/9.6% and 0.0%). This is also shown on table 4b.

**Table 4b:**
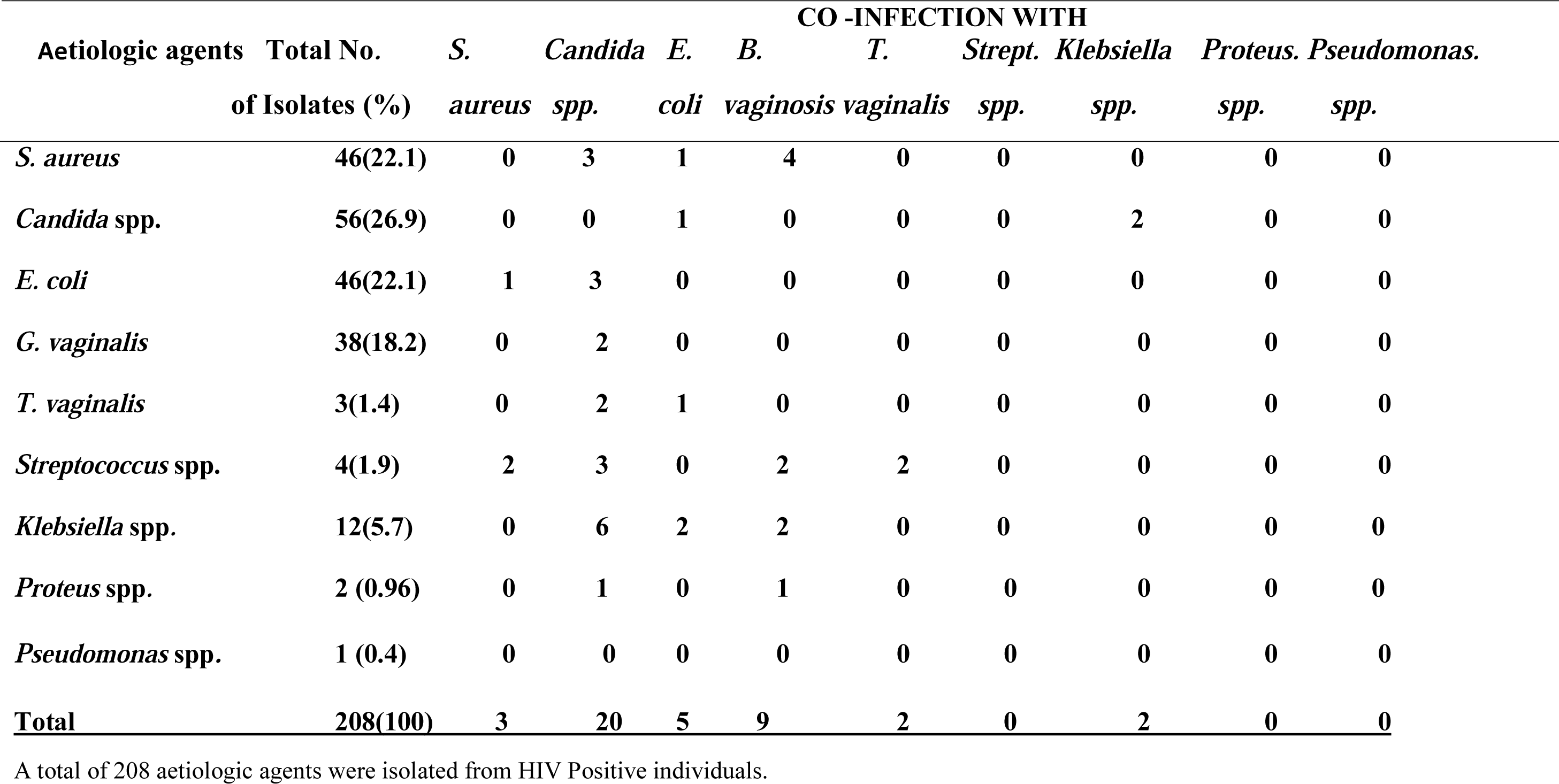
Aetiologic Agents Isolated and Occurrence of Mixed Infections in HIV Positive Individuals

### Long Term Non-progressors (LTNPs)

The display of the length of time the non-progressors have not been on ARV is shown on figure 1. The longest period any of them have lived without drugs is > 7 years (12 HIV positive individuals) and the shortest is 0-12 months (57 HIV positive individuals). Table 5 also shows common documented HLA- A, -B and -C alleles identified among LTNPs of Nigerian origin.

**Figure 1:**
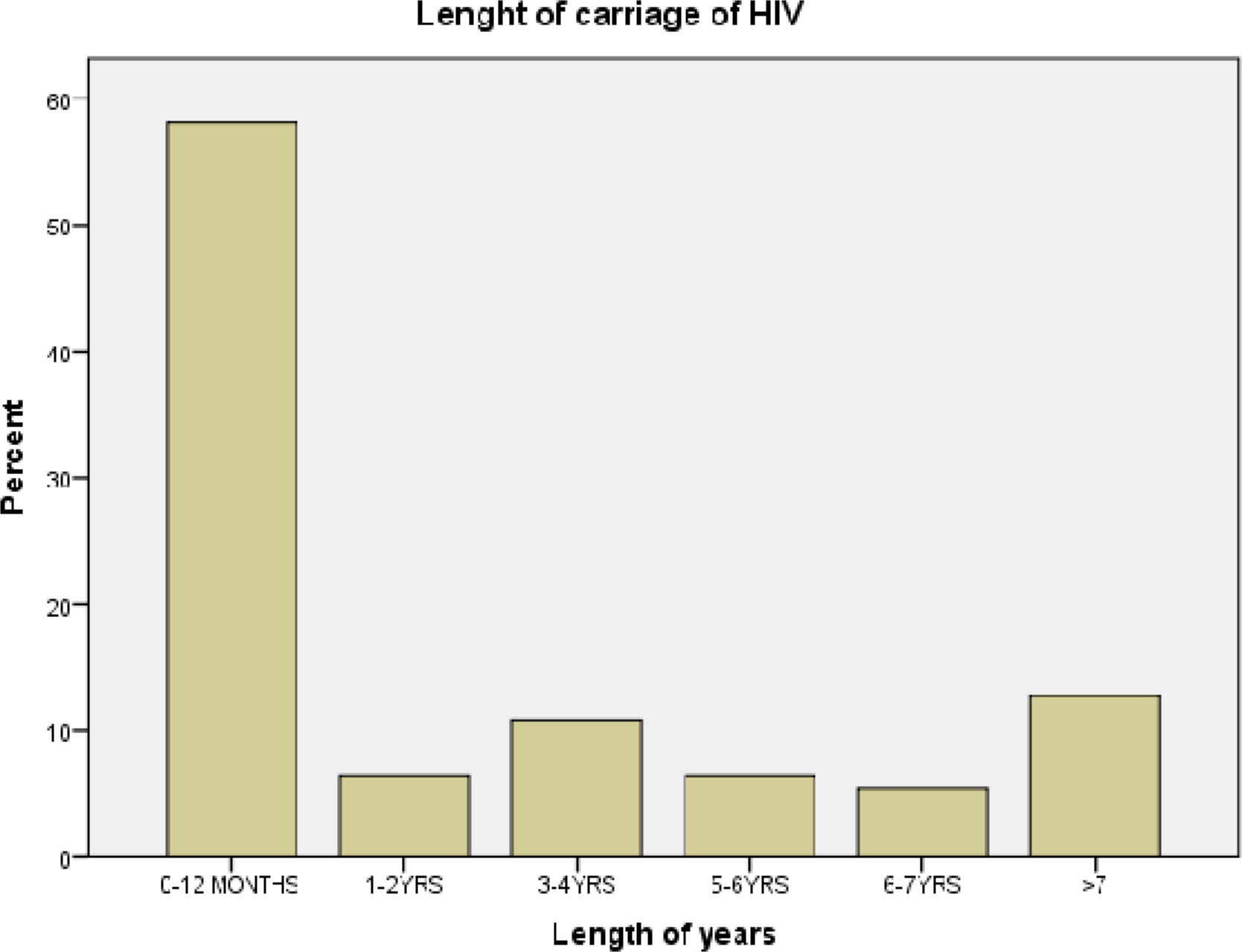
Length of Carriage of HIV in Long-term Non -Progressor’s (LTNP)

**Table 5a:** Antibiotic Susceptibility Pattern of Microbial Agents Isolated from HIV Positive and Negative Individuals

| **HIV** | **Total No** | **Antibiogram Sensitivity Pattern in Percentage (%)** |  |  |  |  |  |  |  |  |  |  |  |  |  |  |  |
| --- | --- | --- | --- | --- | --- | --- | --- | --- | --- | --- | --- | --- | --- | --- | --- | --- | --- |
|  | **of Isolates** |  |  |  |  |  |  |  |  |  |  |  |  |  |  |  |  |
| **Status** | **(%)** | **CXM** | **TET** | **OFL** | **GEN** | **AMOX** | **AUG** | **CPR** | **CTR** | **ERY** | **NIT** | **NAL** | **CAZ** | **CRX** | **CXC** | **COT** | **CTR** |
| **Pos. *S. aureus*** | **22.1** | **0.0** | **13.5** | **45.2** | **90.4** | **9.4** | **95.4** | **13.5** | **22.6** | **54.2** | **31.6** | **0.0** | **13.5** | **76.9** | **31.6** | **0.0** | **0.0** |
| **Neg. *S. aureus*** | **14.5** | **0.0** | **0.0** | **48.2** | **34.4** | **0.0** | **55.1** | **0.0** | **27.5** | **34.4** | **13.2** | **0.0** | **20.6** | **0.0** | **41.6** | **0.0** | **0.0** |
| **Pos. *E. coli*** | **22.1** | **0.0** | **18.1** | **95.4** | **98.0** | **13.5** | **66.3** | **31.6** | **0.0** | **0.0** | **98.5** | **22.6** | **31.6** | **13.5** | **0.0** | **0.0** | **0.0** |
| **Neg. *E. coli*** | **27.2** | **25.3** | **11.0** | **33.0** | **73.5** | **14.7** | **14.7** | **29.4** | **0.0** | **0.0** | **66.1** | **0.0** | **20.4** | **7.35** | **0.0** | **7.35** | **11.0** |
| **Pos. *Strep. spp.*** | **1.9** | **0.0** | **0.0** | **0.0** | **0.0** | **0.0** | **0.0** | **0.0** | **0.0** | **0.0** | **0.0** | **0.0** | **47.5** | **47.5** | **0.0** | **0.0** | **0.0** |
| **Neg. *Strep. spp.*** | **5.5** | **0.0** | **18.1** | **18.1** | **0.0** | **0.0** | **0.0** | **0.0** | **0.0** | **0.0** | **0.0** | **0.0** | **0.0** | **42.3** | **0.0** | **0.0** | **0.0** |
| **Pos. *Kleb. spp*** | **5.7** | **17.5** | **0.0** | **87.7** | **52.6** | **0.0** | **17.5** | **52.5** | **0.0** | **0.0** | **87.7** | **52.6** | **17.5** | **17.5** | **0.0** | **0.0** | **0.0** |
| **Neg. *Kleb. Spp.*** | **7.2** | **0.0** | **27.7** | **27.7** | **0.0** | **0.0** | **27.7** | **0.0** | **0.0** | **0.0** | **0.0** | **27.7** | **27.7** | **13.8** | **41.6** | **13.8** | **0.0** |
| **Pos. *Proteus spp.*** | **0.96** | **20.8** | **0.0** | **32.0** | **32.0** | **0.0** | **20.8** | **32.0** | **0.0** | **0.0** | **10.4** | **0.0** | **10.4** | **10.4** | **0.0** | **0.0** | **0.0** |
| **Neg. *Proteus spp.*** | **3.6** | **0.0** | **0.0** | **27.7** | **27.7** | **0.0** | **0.0** | **27.7** | **0.0** | **0.0** | **27.1** | **0.0** | **0.0** | **0.0** | **0.0** | **0.0** | **0.0** |
| **Pos. *Pseudo. spp.*** | **0.4** | **40** | **0.0** | **40.0** | **40.0** | **0.0** | **0.0** | **0.0** | **0.0** | **40.0** | **20.0** | **20.0** | **20.0** | **0.0** | **0.0** | **0.0** | **0.0** |
| **Pos. *Pseudo. spp.*** | **0.4** | **0.0** | **10.0** | **8.0** | **5.7** | **40.0** | **13.3** | **0.0** | **0.0** | **0.0** | **40.0** | **4.0** | **0.0** | **0.0** | **0.0** | **20.0** | **0.0** |

Pos. is HIV positive status, Neg. is HIV negative status.
0.0 - means the isolates showed no sensitivity to antibiogram

**Table 5b:** Antibiotic Resistant Pattern of Aetiologic agents Isolated from HIV Positive and Negative Individuals

| HIV | Total No | Antibiogram Resistance in Percentage (%) |  |  |  |  |  |  |  |  |  |  |  |  |  |  |  |  |
| --- | --- | --- | --- | --- | --- | --- | --- | --- | --- | --- | --- | --- | --- | --- | --- | --- | --- | --- |
| Aetiologic | of Isolates |  |  |  |  |  |  |  |  |  |  |  |  |  |  |  |  |  |
| Status | Agents | (%) | CXM | TET | OFL | GEN | AMOX | AUG | CPR | CTR | ERY | NIT | NAL | CAZ | CRX | CXC | COT | CTR |
| Pos. *S. aureus* |  | 22.1 | 0.0 | 40.7 | 67.8 | 72.3 | 54.2 | 40.7 | 0.0 | 63.3 | 58.8 | 40.7 | 36.1 | 81.4 | 18.0 | 40.0 | 63.3 | 22.6 |
| Neg. *S. aureus* |  | 14.5 | 0.0 | 68.9 | 62.0 | 6.8 | 13.6 | 48.2 | 6.8 | 27.2 | 55.1 | 6.8 | 16.8 | 68.9 | 55.1 | 48.2 | 6.8 | 13.6 |
| Pos. *E. coli* |  | 22.1 | 9.0 | 85.9 | 76.9 | 36.1 | 63.1 | 66.9 | 4.5 | 0.0 | 9.4 | 58.8 | 72.3 | 36.1 | 27.1 | 3.0 | 73.6 | 0.0 |
| Neg. *E. coli* |  | 27.2 | 18.3 | 11.0 | 33.0 | 18.3 | 25.7 | 44.1 | 14.7 | 3.6 | 11.0 | 25.7 | 7.2 | 29.4 | 25.7 | 3.6 | 11.0 | 7.2 |
| Pos. *Strep. spp.* |  | 1.9 | 0.0 | 52.6 | 95.0 | 95.0 | 0.0 | 95.0 | 0.0 | 95.0 | 0.0 | 95.0 | 95.0 | 95.0 | 0.0 | 52.6 | 0.0 | 0.0 |
| Neg. *Strep. spp.* |  | 5.5 | 0.0 | 18.1 | 81.1 | 54.5 | 0.0 | 54.0 | 0.0 | 81.1 | 0.0 | 0.0 | 18.1 | 0.0 | 0.0 | 18.1 | 18.1 | 18.1 |
| Pos. *Kleb. spp.* |  | 5.7 | 0.0 | 52.6 | 35.5 | 52.6 | 70.1 | 52.6 | 17.5 | 35.3 | 35.3 | 35.3 | 35.3 | 52.6 | 7.5 | 36.3 | 55.3 | 0.0 |
| Neg. *Kleb. spp.* |  | 7.2 | 0.0 | 13.8 | 27.7 | 41.6 | 13.8 | 41.61 | 13.8 | 0.0 | 0.0 | 13.8 | 0.0 | 27.7 | 13.8 | 0.0 | 13.8 | 0.0 |
| Pos. *Proteus spp.* |  | 0.96 | 0.0 | 32.0 | 0.0 | 0.0 | 48.0 | 9.6 | 0.0 | 9.6 | 9.6 | 0.0 | 9.6 | 0.0 | 0.0 | 0.0 | 0.0 | 0.0 |
| Neg. *Proteus spp.* |  | 3.6 | 0.0 | 55.5 | 55.5 | 83.3 | 83.3 | 83.3 | 27.7 | 0.0 | 0.0 | 27.7 | 27.7 | 83.3 | 55.5 | 0.0 | 0.0 | 0.0 |
| Pos. *Pseudo. spp.* |  | 0.4 | 0.0 | 2.0 | 40.0 | 40.0 | 20.0 | 20.0 | 0.0 | 40.0 | 0.0 | 40.0 | 40.0 | 0.0 | 0.0 | 0.0 | 0.0 | 0.0 |
| Neg. *Pseudo. spp.* |  | 0.4 | 0.0 | 2.0 | 13.3 | 20.0 | 10.0 | 40.0 | 0.0 | 0.0 | 0.0 | 0.0 | 0.0 | 0.0 | 0.0 | 0.0 | 40.0 | 0.0 |

Pos. is HIV Positive status and Neg. is HIV negative status.
0.0 - means the isolates showed no resistance to antibiogram

## Discussion

HIV infected subjects acquire new sexually transmitted diseases and reproductive tract infections (STDs/RTIs) at a rate equal to or greater than the HIV negative subjects. This is because these group of people are immunocompromised and are involved in other risky sexual practices such as: failure to use condom during sexual activities, anal or oral sex and some of the patients are involved in poor Antiretroviral drugs (ARV) adherence which may result in HIV mutation and leads to increase in opportunistic infections. Sixty-one (61) index whose HIV-1 plasma viral load (HIV-1 RNA) was categorized at baseline: <1000 to ≥ 1,000,000copies/ml, showed a significant association between baseline HIV-1 plasma viral load, heterosexual HIV transmission (p=0.059) and STIs/RTIs (p=0.057). Again, inheriting the HLA susceptible allele C*07:02, B*07:02 and A*23:01 with increased plasma viral load, presence of untreated STIs, non-use of condom has been established from previous studies, to expose the sexual partners to high risk of HIV acquisition^22^.

In this study, a total of 208 (*p*=0.05) microorganisms were isolated from HIV positive individuals compared to 55 isolated from HIV negative individuals. *Candida* spp. 56 (26.9%) was the most prevalent isolated from HIV positive individuals followed by *Staphylococcus aureus and Escherichia coli* 46 (22.1%) respectively. This result is consistent with a study done among people living with HIV/AIDs in Port Harcourt^41^. In this report, *Staphylococcus aureus* had the highest percentage of occurrence 49 (29.7%), followed by *Escherichia coli* 47 (28.5%), *Pseudomonas aeruginosa* 46 (27.9%) and *Klebsiella pneumoniae* 23 (13.9%). Most of these organisms especially *Candida* spp. (26.9%) *and Gardnerella vaginalis* (18.2%) were opportunistic infections occurring in immunocompromised women. Immunocompromised HIV-1 infected women are known to have increased rates of vaginal candidiasis with increased severity of pelvic inflammatory diseases.

This result is also consistent with another study done on lower genital tract infections in HIV-seropositive women in India^9^. The laboratory findings showed high prevalence of BV (30%), mixed infection (30%), and candidiasis (10%) among HIV-seropositive women (*p* <0.001). But contrasts with other findings in the State of São Paulo^42^. In their study, they reported that *Pseudomonas aeruginosa* was the most frequently isolated microorganism, followed by *Klebsiella pneumoniae* in AIDs patients. In this study however, *Escherichia coli* and *Staphylococcus aureus* (22.1%) respectively were the most frequently isolated microorganism followed by *Klebsiella* spp. 12(5.7%) while *Pseudomonas* spp. was the least frequent 1(0.4%). Again, *Candida* spp. and *Gardnerella vaginalis* co-infected mostly with *Klebsiella* spp. (6/2 times) consecutively.

Out of the bacterial isolates that exhibited multidrug resistance, *Staphylococcus aureus* and *Escherichia coli* isolated from HIV positive individuals showed more resistance to Ceftazidime, (81.4%) Gentamycin (72.3%) and Tetracycline (85.9%). This result is comparable to a recent study that reported that *Staphylococcus aureus* isolated from HIV positive individuals recorded the highest number of multidrug resistant bacteria 36 (32.4%)^41^. The high levels of multidrug resistance in HIV seropositive individuals are a serious public Health concern in low income countries. This is because there is a lot of across- the counter purchase of antibiotics without doctor’s prescription or laboratory test results. Result from the questionnaire shows that 70% of the clients who had itching, painful urination, vaginal and urethral discharge have had self-medication of antibiotics mostly of substandard quality and only reports to the hospital when they cannot manage the situation. This is due to ignorance, poverty, poor hygiene practices as well as inconsistent use of condom.

From previous studies, different HLA class I alleles have been significantly associated with different rates of HIV diseases progression to AIDS while some haplotypes have been responsible for HIV-1 viral control during the last stage of HIV infection^9^. The HLA class I profile of Long-term Non-Progressors (LTNP) among the index (HIV positive subjects) were reviewed along baseline CD4+ count and plasma viral load (HIV-1 RNA). This was aimed to identify and match those alleles that have been described in other HLA studies in other African, African American countries as being independently associated with HIV disease protection, susceptibility and or alleles influencing seroconversion. In this study, it was observed that this group of indexes have HLA class I genes associated with low plasma viral loads (HIV-1 RNA) and high CD4+counts (highest/lowest CD4+ counts were 920/402cells/mm3 while the highest/lowest pVL were 181/20 copies/ml). Twenty-four, 24 (8.51%) out of 282(54.02%) were LTNPs. However, 80% (P=0.05) of the partners of LTNP were HIV negative while 20% were HIV Positive. HIV-1 plasma viral loads can independently or in association with different HLA allele allotypes may contribute to HIV-1 transmission among Index-donor and Partner-recipient pairs. A study reported that any reduction in the plasma HIV-1 RNA is an important target for HIV prevention^15^. Another study of heterosexual Uganda HIV serodiscordant couples showed an association between the risk of HIV acquisition and donor plasma viral load (pVL). In their report, there was no HIV transmission observed when the pVL was below the threshold of 1,500 copies HIV-1 RNA/ml^43^ which is consistent with the 80%, in this study whose partners were HIV-1 seronegative. However, we identified 20% of the partners of the index to be HIV-1 seropositive though the index highest pVL recorded was 181copies/ml. However, this result is consistent with a recent study^44^ who estimated the risk of HIV-1 acquisition in a sole sexual exposure estimated to be very low if the host viral load is low. It is well understood that one of the significant factors that affects the heterosexual transmission of HIV-1 in serodiscordant couples is the exposing dose of the virus in the plasma of the index. However, further studies may be able to identify why 20% of the partners of the index were seropositive when the highest pVL was below 200copies/ml.

In this study also, 3(12.5%) of LTNPs inherited HLA B*57:01(12.9%) and B*57:03 (4.6%) which are haplotypes known to be responsible for HIV-1 viral control during the last stage of infection from previous studies^9^. HLA-B* 57:03 (4.6%) was also documented to be responsible for slowing CD4+ decline. The beneficial HLA alleles *A1 (4.16%), C*07:01(29.1%) and C*06:02 (12.5%) were identified as alleles independently associated with protection from seroconversion. However, B*07:02/C*07:02 (33.3%) and C*23:01(4.16%) which was shown in previous studies^22^ to be independently associated with seroconversion to HIV diseases was also identified within the cohort of LTNPs. It is therefore suggested that the LTNPs in this cohort may be carriers of non-virulent type of HIV or that their HLA generates an immunodominant response to an epitope in the Gag protein 24 and dynamic CD8+ responses which is critical for long- term control of HIV replication in these individuals. However, any escape from this response may lead to significant HIV seroconversion, increased viral loads and reduced CD4+ counts. Another study reported that HLA alleles that is defending the immune system have a true preference for the p24 Gag protein and CTL responses against Gag which moderately reduces HIV disease evolution^16^. Twelve percent (12%) of the LTNPs in this study have known their HIV status over 7 years while 58% have known their HIV-1 status between 0-12 months. Another study reported that 11 of the 13 LTNP 85%) had a gene encoding HLA- B*57:01 variant which has been implicated to slow HIV disease evolution^45^. This study observed that LTNPs have HLA-B*57:01/*5703(17.5%) and other documented common alleles. Again, LTNPs HLA-C alleles may be implicated with increased surface expression which is associated with reduced viral load and improved CD4^+^ T cell counts. This High HLA-C expression have improved HIV control through active cytotoxic CD8^+^ T cell responses as recorded in the Nigerians, African, European Americans^46, 47^.

## Conclusion

Microorganisms isolated from the index were associated with high viral loads leading to low immunity and so are independent makers to HIV-1 transmission among serodiscordant couples. There is need to intensify health education to create public awareness on the use of low standard antibiotics purchased across the counter and self- medication. This will reduce multi drug resistance and prevent reemergence of opportunistic infections in the index. HLA class I alleles identified among LTNPs were similar to those significantly associated with resistance and susceptible to HIV-1 disease.

## Data Availability

Data will be made available on request.

## Acknowledgement

I deeply appreciate HIV Research Trust Scholarship UK 2012 (HIVRT12-082) for sponsoring the preliminary training I required for this project which included, traveling to Project San Francisco (PSF) Kigali, Rwanda and Zambia Emory HIV Research Project (ZEHRP) Lusaka to get trained on Couples HIV Voluntary Counselling and testing, white cell pellet harvesting, DNA extraction and PCR. This was done under the supervision of Prof Susan Allen. I am grateful to Dr. Ma Lou. Associate Professor, University of Manitoba, HIV Host Genetics, National Microbiology Laboratories Winnipeg Canada for the provision of reagent and equipment used for the HLA studies as well as the analysis. My warm appreciation goes to Prof. Babatunde Salako, the Director General of the Nigerian Institute of Medical Research, who approved my Pre-Doctoral Research Development Grant through its’ Graduate Initiative Programme (GIP) NIMR which was used for the completion of this study.

## Duality of interest

The authors have declared no duality of interests

## Notes

### Competing Interest Statement

The authors have declared no competing interest.

### Funding Statement

Supported by Pre-Doctoral Research Development Grant from The Nigerian Institute of Medical Research and HIV Research Trust Scholarship UK 2012 (HIVRT12-082)

### Author Declarations

Approval was obtained from NIMR IRB (IRB/12/176) and NAUTH Ethical Committee (CS/66/7/79).

